# Generative Modeling of the Circle of Willis Using 3D-StyleGAN

**DOI:** 10.1101/2024.04.02.24305197

**Authors:** Orhun Utku Aydin, Adam Hilbert, Alexander Koch, Felix Lohrke, Jana Rieger, Satoru Tanioka, Dietmar Frey

**Affiliations:** CLAIM - Charite Lab for Artificial Intelligence in Medicine, Charite Universitatsmedizin Berlin, Germany

**Keywords:** Style-GAN, generative AI, generative modeling, vessel segmentation, TOF MRA

## Abstract

The circle of Willis (CoW) is a network of cerebral arteries with significant inter-individual anatomical variations. Deep learning has been used to characterize and quantify the status of the CoW in various applications for the diagnosis and treatment of cerebrovascular disease. In medical imaging, the performance of deep learning models is limited by the diversity and size of training datasets. To address medical data scarcity, generative adversarial networks (GANs) have been applied to generate synthetic vessel neuroimaging data. However, the proposed methods produce synthetic data with limited anatomical fidelity or downstream utility in tasks concerning vessel characteristics.

We adapted the StyleGANv2 architecture to 3D to synthesize Time-of-Flight Magnetic Resonance Angiography (TOF MRA) volumes of the CoW. For generative modeling, we used 1782 individual TOF MRA scans from 6 open source datasets. To train the adapted 3D StyleGAN model with limited data we employed differentiable data augmentations and used mixed precision and a cropped region of interest of size 32×128×128 to tackle computational constraints. The performance was evaluated quantitatively using the Fréchet Inception Distance (FID), MedicalNet distance (MD) and Area Under the Curve of the Precision and Recall Curve for Distributions (AUC-PRD). Qualitative analysis was performed via a visual Turing test. We demonstrated the utility of generated data in a downstream task of multiclass semantic segmentation of CoW arteries. Vessel segmentation performance was assessed quantitatively using the Dice coefficient and the Hausdorff distance.

The best-performing 3D StyleGANv2 architecture generated high-quality and diverse synthetic TOF MRA volumes (FID: 12.17, MD: 0.00078, AUC-PRD: 0.9610). Multiclass vessel segmentation models trained on synthetic data alone achieved comparable performance to models trained using real data in most arteries.

In conclusion, generative modeling of the Circle of Willis via synthesis of 3D TOF MRA data paves the way for generalizable deep learning applications in cerebrovascular disease. In the future, the extensions of the provided methodology to other medical imaging problems or modalities with the inclusion of pathological datasets has the potential to advance the development of more robust models for clinical applications.

## Introduction

The Circle of Willis (CoW) is a network of cerebral arteries in the skull base and provides collateral blood supply to the brain. CoW has been a significant region of interest in medical image analysis due to pathologies arising in the anatomical proximity and its potential to be used as a biomarker for cerebrovascular disease risk and prognosis (Gutierrez et al., 2015; van Seeters et al., 2015). Several medical interventions and surgeries are performed via the CoW in patients with ischemic stroke (mechanical thrombectomy), cerebral aneurysm (coiling, clipping, flow diverters) and vascular malformations. CoW is an anatomical landmark that exhibits high inter-individual anatomical variations including hypoplasia or aplasia of (communicating) artery segments, and diverse branching patterns of the middle cerebral artery (Iqbal, 2013; Krabbe-Hartkamp et al., 1998). These variations are of vital importance for individualized risk assessment and treatment planning. Therefore, analysis of CoW anatomy and pathologies is regularly performed in the clinical setting as part of imaging protocols based on computed tomography angiography (CTA) and magnetic resonance angiography (MRA) scans.

Deep learning has been used to characterize and quantify the status of the CoW for various applications in stroke including large vessel occlusion (LVO) detection (Brugnara et al., 2023), collateral status assessment (Bagcilar et al., 2023; Grunwald et al., 2019), hemodynamic modeling (Frey et al., 2021) and multiclass semantic segmentation (Yang et al., 2024). The performance and generalization of deep learning models for these tasks significantly depend on the diversity, quality and size of the training dataset. This especially applies to datasets with class imbalance due to rare diseases or underrepresented anatomical variations. Variations of the CoW are vast and vessel neuroimaging data is limited, costly to acquire and difficult to share due to data protection regulations. Previous research has reported that rare variations of the CoW can limit the performance in multiclass anatomical segmentation of CoW arteries (Hilbert et al., 2022; Yang et al., 2024). Several studies report that datasets used for AI-based medical imaging applications suffer from a diversity problem in various tasks (Arora et al., 2023; Hofmanninger et al., 2020). Generative adversarial networks (GANs), a subtype of deep learning methods, enable the synthesis of high-quality, diverse synthetic data representative of the real-world data distribution of the training dataset. They have demonstrated their potential to capture and model inter-individual anatomical and pathological variabilities as well as variabilities arising from different scanner hardware and choice of imaging parameters. Synthetic data for training of downstream deep learning methods in medical imaging has been shown to improve performance in the classification of histology slides (Chen et al., 2021) and image segmentation tasks (Liu et al., 2023). However, current generative models for synthesizing angiography scans such as time of flight magnetic resonance angiography (TOF MRA) of cerebral arteries are based on either 2D data (Kossen et al., 2021) or do not preserve anatomical fidelity and continuity (Subramaniam et al., 2022). While 2D- or 3D-patch-based approaches showcase significant utility for selected downstream tasks, particular downstream tasks such as hemodynamic modeling (Rundfeldt et al., 2024) or 3D classification demand realistic anatomical accuracy and spatial continuity from the synthesized vessel volumes. To tackle the research gap of 3D modeling, prior works have proposed 3D adaptations of generative adversarial networks (Hong et al., 2021; Mensing et al., 2022) and denoising diffusion probabilistic models for medical images (Khader et al., 2023).

In this work we set out to model the complex anatomy and heterogeneous inter-individual variations of the CoW using generative adversarial networks. We adapt the StyleGANv2 architecture (Karras et al., 2020b) to process three-dimensional medical data in clinically relevant high resolutions. We evaluate the influence of model complexity and differentiable data augmentation (Zhao et al., 2020) on generated image quality and diversity. We showcase the capabilities of StyleGANv2 by introducing stochastic variations to the generated TOF MRA images, exploring the latent space of the model with image interpolations and controlling the diversity of the images using the truncation trick. Finally, we validate the generation quality and diversity of the generated images using quantitative metrics and showcase the utility of the model in the downstream task of multiclass semantic segmentation of CoW artery segments. Leveraging concepts from knowledge distillation and self-training, we employ a novel downstream validation cycle where we (1) train a vessel segmentation model on real data, (2) use it to generate labels on synthetic data, (3) use the resulting synthetic data-label pairs to train a new model which we evaluate on real unseen test data.

## Methods

### StyleGAN Model

An open-source implementation of the 2D StyleGANv2 architecture (Karras et al., 2020b) was used as a baseline (Wang, 2024). The architecture was adapted by replacing all upsampling and convolutional operations to 3D similar to a prior 3D StyleGANv2 implementation (Hong et al., 2021). An overview of the generator and discriminator architectures are presented in Figure 1.

**Figure 1.**
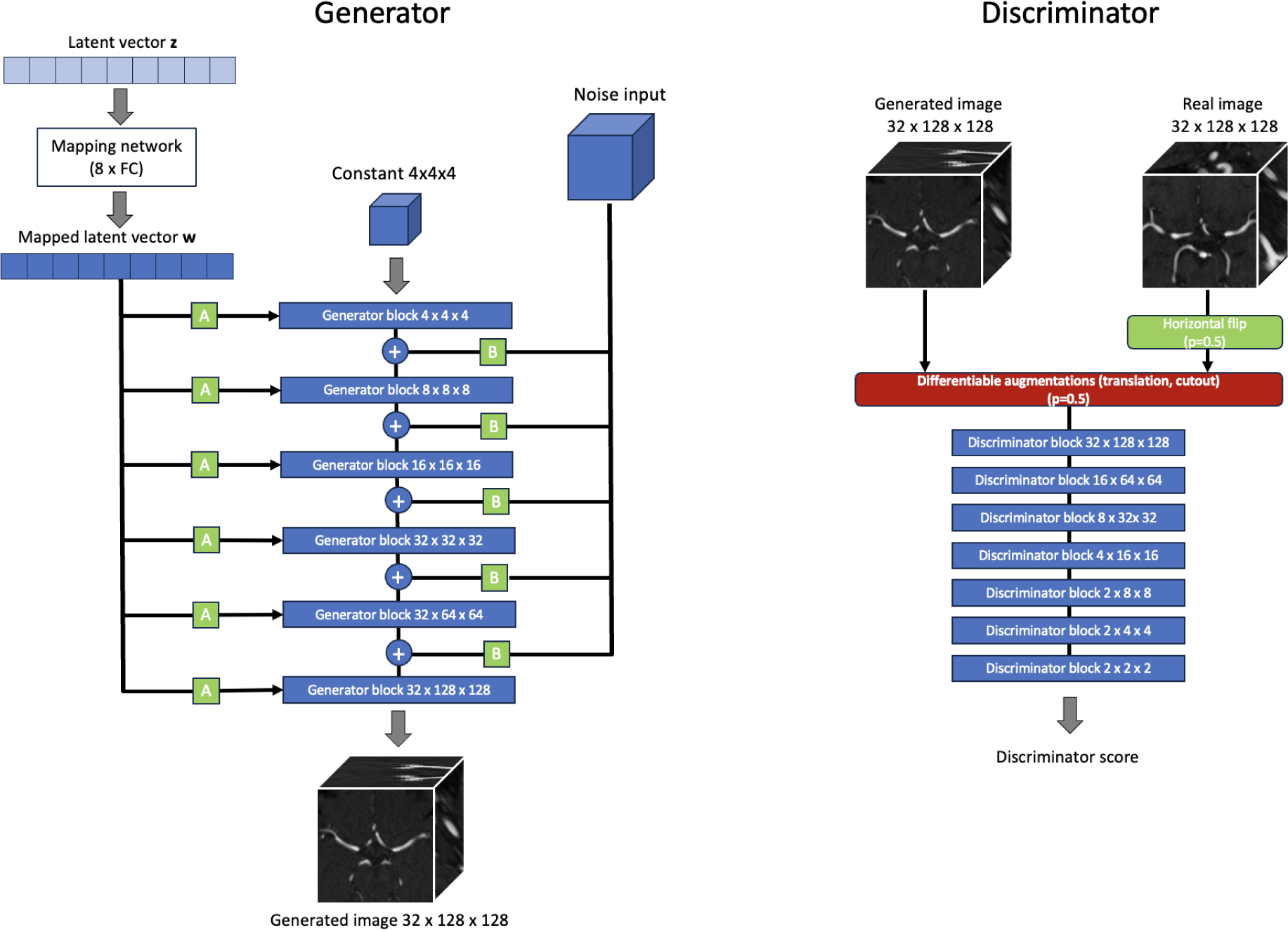
Simplified visualization of the StyleGAN2 architecture adapted for 3D medical images. The two inputs to the generator are 1) the latent vector z for the style vector and 2) noise input that is added after each Generator block. “A” represents a learned affine transformation that converts w to a style vector, “B” defines a noise broadcasting operation. All images that the discriminator sees are augmented with differentiable augmentations with a probability of 0.5. Details of the generator and discriminator blocks, upsampling, flattening and classification layers are omitted for visual clarity. A sample input size of 32×128×128 was selected for visualization.

### Mapping network

The starting point for generation is a noise vector of length 512 with a standard normal distribution. The noise vector is converted to a style vector via the mapping network. The mapping network is a fully connected neural network with 8 layers each followed by a leaky ReLU activation function. The learning rate multiplier was 0.1 for the mapping network leading to a ten times lower learning rate for the mapping network compared to the generator learning rate.

### Discriminator

The discriminator of the StyleGANv2 is composed of a sequential stack of discriminator blocks (Karras et al., 2020b). The number of blocks is dynamically determined by the input image size. For an input image size of 32×128×128, the discriminator consists of 7 discriminator blocks. This configuration is derived by calculating the base-2 logarithm of the largest dimension of the input image. The discriminator blocks are followed by a final 3D convolutional layer, flattening layer and linear layer. Each discriminator block uses two 3D convolutional layers with leaky ReLU activation functions, and integrates a residual connection that merges the input and output of the convolutional layers. The number of convolutional filters doubles at each block until the maximum set number of filters is reached. The discriminator blocks progressively reduce the spatial resolution of feature maps via strided convolutions and blurring operation across all dimensions until a dimension of 2 is reached, then only remaining dimensions greater than 2 are downsampled.

The loss function of the discriminator consisted of the hinge loss (Lim and Ye, 2017) and the gradient penalty term to maintain the Lipschitz constraint (Gulrajani et al., 2017). We apply lazy regularization every 16 training steps-instead of every step- to save computational resources (Karras et al., 2020b). The loss function equation for the discriminator was:

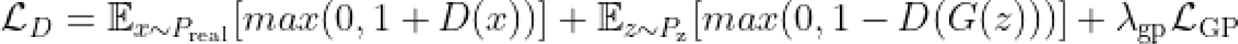

where x is drawn from the distribution of real images P_real_, z is the latent noise vector, D is the discriminator, G is the generator, L_GP_ is the gradient penalty term with weight λ_GP_ of 10.

### Generator

The generator architecture begins with an initial block that generates the first tensor from a constant 4×4×4 tensor. Following the initial block, the architecture has a sequence of generator blocks, each responsible for doubling the resolution of its input tensor. For an input image with dimensions of 32×128×128, the generator is structured to include 6 discriminator blocks. The StyleGANv2 generator architecture incorporates modulated convolutions within its generator blocks (Karras et al., 2020b), where convolutional filters are dynamically adjusted based on input style vectors, followed by a demodulation step to normalize the outputs. Each generator block performs either isotropic trilinear upsampling, where all dimensions are scaled by a factor of 2, or anisotropic trilinear upsampling, which selectively upscales the width and height dimensions, leaving the depth dimension unchanged. Each block uses two convolutional layers modulated by style vectors. After each convolutional layer, a noise input of the same dimensions as the block’s output is added to the feature maps, and a leaky ReLU activation is applied. The introduction of noise at each layer helps to introduce fine-grained details and textures into the generated images, making them more realistic. The loss function of the generator consisted of a generator hinge loss and a path length penalty introduced after 5000 steps every 32 steps during the training. The path length penalty aims to encourage the generator to produce consistent and smooth variations in the output images as the input latent space is changed. The equation for the path length penalty term was:

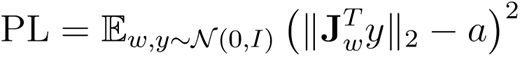

where J_w_ denotes the Jacobian matrix of the generator network’s output with respect to the latent vector w, “y” is a noise vector sampled from the standard normal distribution N(0, I) with the same shape as generators output and “a” the target path length which is a running average of path lengths during training.

The resulting generator loss function was:

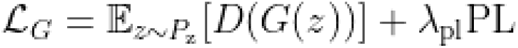

where D is the discriminator, G is the generator and PL denotes the path length penalty term weighted by a regularization coefficient λ_PL_ =1.

### Truncation trick

Truncation of style vectors has been proposed previously to control the trade-off between the fidelity and diversity of generated images (Brock et al., 2019; Karras et al., 2019). This can be achieved by first calculating a mean style vector by passing noise vectors z through the mapping network. Each individual style vector can then be adjusted based on a truncation parameter towards the mean style vector. Lower values of the truncation parameter lead to images that are more closely aligned with the central style, enhancing their realism but potentially at the expense of diversity. The effect of the truncation parameter ψ with respect to quality and diversity was visually analyzed by generating example images with ψ = 1, 0.75, 0.5, 0.25. The truncation trick was implemented as follows:

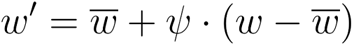

where w’ is the truncated style vector, *w̅* is the average style vector, w is the original style vector before truncation and ψ is the truncation parameter with typical values between 0 and 1.

### Data

Open source TOF MRA data from 7 datasets were used. 1782 scans from 6 datasets were used to train the 3D StyleGAN model. The TopCoW (Topology-Aware Anatomical Segmentation of the Circle of Willis for CTA and MRA) challenge dataset (n=110) was used for validation and the downstream multiclass CoW segmentation analysis (Yang et al., 2024). Importantly, the TopCoW dataset included patients with stroke-related neurological disorders and stood in contrast to the datasets used for the training of the StyleGAN model. In the training set for generative modelling all datasets included healthy patients except for OASIS 3 dataset which also included patients with neurodegenerative disease (LaMontagne et al., 2019). The relevant characteristics of the datasets are described in Table 1. The datasets were acquired using 3T and 1.5 T MRI machines from different vendors. Further descriptions on patient characteristics and imaging protocols can be found in the respective publications.

**Table 1.** Datasets used in the study. A total of 1782 TOF MRA scans were used from 6 open source datasets for generative modeling. For downstream validation, the TopCoW Challenge dataset was used both for training and testing of multiclass CoW artery segmentation models.

| Dataset | Number of scans | Site | Population | Modeling | Citation |
| --- | --- | --- | --- | --- | --- |
| IXI | 564 | Multi | Healthy | Training | ("IXI Dataset – Brain Development," n.d.) |
| Lausanne | 126 | Single | Healthy | Training | (Di Noto et al., 2023) |
| NITRC | 56 | Single | Healthy | Training | ("NITRC: Magnetic Resonance Angiography Atlas Dataset: Tool/Resource Info," n.d.) |
| CASILab | 109 | Single | Healthy | Training | (Bullitt et al., 2005) |
| ICBM | 180 | Multi | Healthy | Training | (Mazziotta et al., 2001) |
| OASIS 3 | 747 | Single | Neurodegenerative disease | Training | (LaMontagne et al., 2019) |
| TopCow | 110 | Multi | Stroke-related neurological disorder | Validation | (Yang et al., 2024) |

### Data preprocessing pipeline

A custom TOF MRA template with a size of (288×320×199 in width,height,depth) voxels and a voxel spacing of 0.62×0.62×0.62 was created using the 56 patients from the NITRC dataset. First, a reference image with no artifacts was selected from the dataset. Each image in the dataset was registered to the reference image using FLIRT (FMRIB’s Linear Image Registration Tool) of FSL (FMRIB Software Library) (Jenkinson et al., 2012). Registered images were averaged using the “fslmaths” command to create the custom template image. Skullstripping of the custom template and training images was performed using SynthStrip (Hoopes et al., 2022). The skullstripped images were registered to the custom template using FLIRT (FMRIB’s Linear Image Registration Tool) with 6 degrees of freedom and mutual information as the cost function (Jenkinson et al., 2002; Jenkinson and Smith, 2001). The resulting transformation matrix was applied to the original images without skull stripping, ensuring that arteries of the skull base are not omitted due to preprocessing errors. Images where any of the preprocessing steps failed were excluded. The preprocessing steps are illustrated in Figure 2.

**Figure 2.**
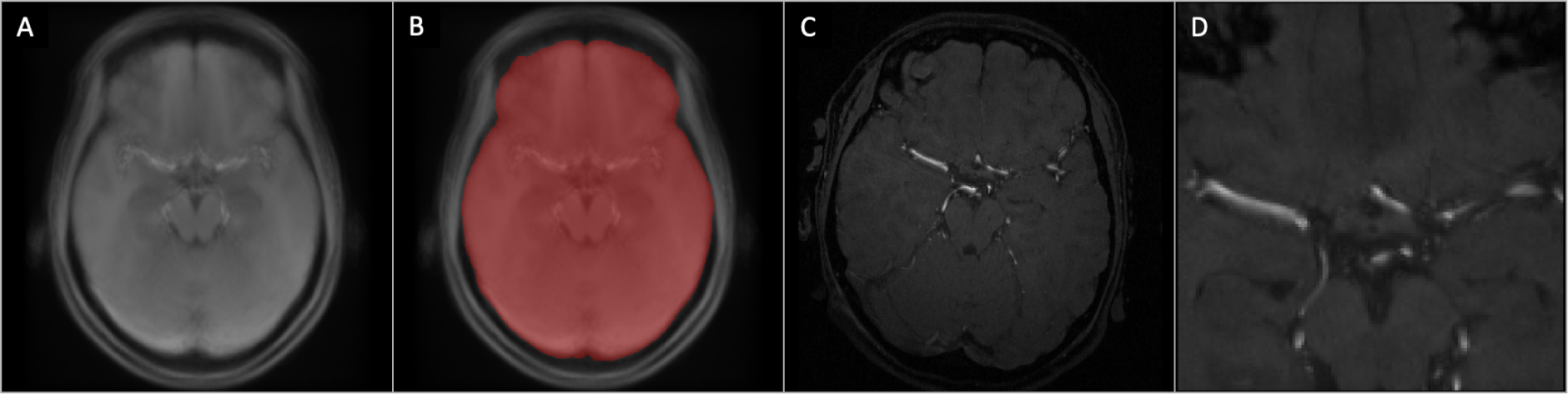
Preprocessing of the real training data. A) Custom TOF MRA template generated from the NITRC dataset. B) Brain mask overlaid on the custom template. Skullstripped images were used for registration and the resulting transformation matrix was applied to non-skullstripped TOF MRAs. C) An axial slice of an example real image. D) The cropped region of interest of size 32×128×128 after registration to the TOF MRA template.

### Model training

StyleGANv2 architectures with varying complexity levels were explored by changing the number of filters for the first layer and the maximum number of convolutional filters (Table 2). All real and generated images seen by the discriminator were augmented using different combinations of translation, cutout and contrast as differentiable augmentations with an augmentation probability of 0.5 (Zhao et al., 2020). Horizontal flipping was applied with a probability of 0.5 on real data to effectively double the dataset size. The Adam optimizer was used with β_1_ = 0. 5 and β_2_ = 0. 9 (Kingma and Ba, 2017). The learning rate was set using a Two Time-scale Update Rule (TTUR) multiplier of 1.5, resulting in initial learning rates of 0.0001 and 0.00015 for the generator and discriminator respectively (Heusel et al., 2017). The learning rate for the configuration *Large* was set lower (G: 0.00005, D:0.000075) to ensure stability. The effective batch size was set to 32 using gradient accumulation 4 times in each step with a batch size of 8. The Config-L was trained with a batch size of 4 and gradient accumulation 8 times for each step, resulting in the same effective batch size of 32.

**Table 2.** Model configurations and hyperparameters.

| Model name | Generator Init Conv Filter | Discriminator Init Conv Filter | Max Conv Filters | Number of parameters (million) Generator / Discriminator |
| --- | --- | --- | --- | --- |
| Small | 16 | 32 | 256 | 12 / 22 |
| Medium_v1 | 16 | 32 | 512 | 30 / 67 |
| Medium_v2 | 16 | 32 | 1024 | 30 / 181 |
| Large | 32 | 64 | 1024 | 117 / 267 |

Each model was trained on a single V100 GPU with 32 GB VRAM in a high performance research cluster. Mixed precision (Micikevicius et al., 2018) was implemented using the automated mixed precision (AMP) functionality in PyTorch to reduce memory footprint and speed-up the training.

## Evaluation

### Stochastic variation

Medical images contain stochastic elements due to image noise, scanner variations or patient movement. When synthesizing artificial medical images, this is a key characteristic that should be preserved to create realistic diversity in the synthetic dataset. These stochastic variations can be modelled by changing the random noise input of the StyleGAN model while keeping the style vector constant leading to generated images with similar anatomical structure but slight changes in random aspects of the image. Stochastic variations were demonstrated by generating 100 images from the same style vector varying only the additional noise input. The coefficient of variation per pixel across 100 volumes was calculated and axial slices were visualized to demonstrate the locations most affected by changes to the noise input.

### Style Mixing

Style mixing enables style-based generative models to generate samples by inheriting aspects from two distinct style vectors. Style mixing can be used to combine different anatomical and imaging characteristics, demonstrating the ability of the generative model to reproduce the wide range of anatomical features present in different subjects. To explore the model’s ability to smoothly transition between these diverse features, spherical linear interpolation (Shoemake, 1985; White, 2016) was used in the latent space. A series of TOF MRA volumes were generated by interpolating in 20 steps between two latent vectors. By generating a TOF MRA volume using each intermediate latent vector while moving from one latent vector to another the transition from one generated image to another was visualized. The interpolations were performed at the level of the latent noise vector before passing through the mapping network.

### Evaluation of fidelity and diversity

Quantitative evaluation was performed using 3 metrics: Fréchet Inception Distance (FID) (Heusel et al., 2017), MedicalNet Distance (MD) (Chen et al., 2019), and area under the curve of the Precision Recall for Distribution (AUC-PRD) (Sajjadi et al., 2018). The Fréchet Inception Distance was computed slice-wise using all 32 axial slices of real and generated 3D volumes. An additional metric based on the Fréchet Distance was computed using the MedicalNet (Chen et al., 2019) as the feature extractor and is referred to as MedicalNet Distance (MD) in this manuscript. This follows prior works (Subramaniam et al., 2022; Sun et al., 2022) and has the goal to compute a domain adapted version of the FID. The AUC-PRD (Sajjadi et al., 2018) was computed based on the extracted features by the MedicalNet to assess the diversity (recall) and fidelity (precision) of the generated images in a single value. The FID, MD and the AUC-PRD scores were monitored during training every 3000 steps. The metrics were also computed using two random subsets of the real training data to get a benchmark of expected values.

Qualitative visual analysis was performed throughout training and testing of models. Additionally, a visual Turing test was performed using 100 TOF MRA volumes (50 generated and 50 real volumes). The best performing model based on the quantitative metrics was used to generate the volumes. The bottom and top 2 axial slices were removed from the volumes before visual rating, since these slices most frequently contained artifacts introduced by the generative model. The visual rating was performed by a board-certified stroke specialist (ST) with more than 10 years of experience. The rater assessed the TOF MRA volumes one-by-one in 3D using ITK-SNAP (Yushkevich et al., 2016), was blinded to the origin of the images and had 1 minute to classify each volume as either real or generated.

### Replica Detection

Generative AI studies in medicine report generated images that are copies of the original training data (Dar et al., 2024). It is important to ensure that the generated data is unique and the generative model does not memorize training data and simply produce identical copies from the training dataset. A replica detection method based on the L2 distance between real and generated images was used (Carlini et al., 2023; Yoon et al., 2023). The criterion for a generated image to be considered a replica was that the closest real image as measured by L2 distance should have an L2 distance that is less than ⅓ of the L2 distance of the second closest real image (Yoon et al., 2023). This indicates that a generated image is unusually similar to a real image, potentially indicating a replica. The formula for the L2 distance based criterion for replica detection was:

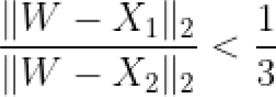

where W is a generated sample, X_1_, X_2_ are the samples with the smallest and second smallest L2 distance to W.

We report the percentage of suspected replicas via the L2 distance methodology using 500 generated TOF MRA volumes and visually verify the authenticity of a subset of 50 random generated images by checking the visual similarity to the corresponding real images with the closest L2 distance.

### Downstream multiclass CoW artery segmentation

Multiclass segmentation of CoW arteries was used as a downstream task to show the utility of the generated data. The TopCow challenge data was split into a training (50) and test set (60). For paired artery segments the left and right labels were merged to create a single class. This resulted in following artery segments as target classes: Internal carotid artery (ICA), basilar artery (BA), posterior communicating artery (Pcom), anterior communicating artery (Acom), the posterior cerebral artery (PCA), anterior cerebral artery (ACA) and the first segment of the middle cerebral artery (M1). The same preprocessing as in the training dataset was applied on the provided full TOF MRA scans, including rigid registration and center cropping. For CoW artery segmentation, the nnUnet framework was used which is a self-configuring state of the art medical image segmentation method (Isensee et al., 2021). A default 3D nnUnet model was trained on the training set of 50 TOF images of real patients using 5-fold cross validation. We refer to this model as the teacher nnUnet model. The performance of the model was assessed on the test set consisting of 60 real patients.

Prior works have proposed to generate the labels together with the original images (Li et al., 2022; Subramaniam et al., 2022). This can be achieved by concatenating labels to the original images as a second channel. While this is practical for downstream model training, it increases the memory footprint of the models due to the added channel. We hypothesize, under the assumption that the generated images are of high enough quality, that a model trained on real data should be able to produce high quality labels when inference is performed on generated data. Therefore, we formulate a validation cycle for our models visualized in Figure 3. First, as an indirect validation of the quality of generated images, the teacher nnUnet model trained on real data was used to perform vessel segmentations on the generated data. Second, the segmentations provided by the teacher model were used as pseudo-labels, i.e. a silver-standard, to train a student nnUnet model with identical hyperparameters and same 5-fold cross validation as the teacher model on synthetic datasets of varying size and diversity. Finally, the performance of the student model was evaluated on the real test data using the Dice coefficient (Zou et al., 2004) and Hausdorff Distance (Taha and Hanbury, 2015) and compared against the teacher model using the EvaluateSegmentation tool (Taha and Hanbury, 2015).

**Figure 3.**
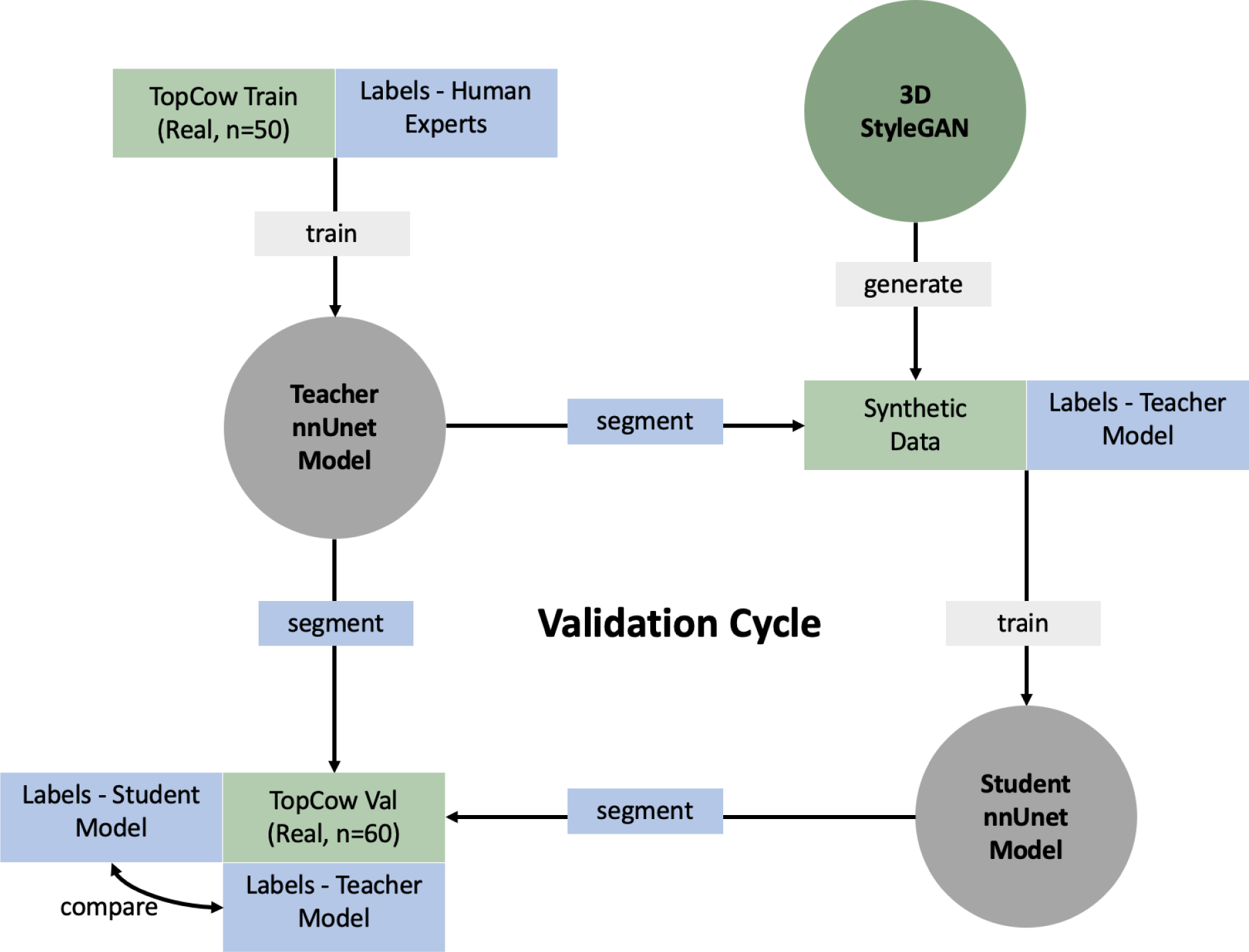
Concept of training a student segmentation model on synthetic data and labels provided by the teacher segmentation model. The quality of the synthetic data is validated by 1) the ability to segment synthetic data with a model trained on real data, 2) training a student model on synthetic data that can segment arteries of the Circle of WIllis on real test data. For quantitative evaluation of segmentation performance the labels provided by the teacher and student models were compared using the TopCow Validation set of 60 patients.

## Results

The proposed 3D adaptation of the StyleGANv2 architecture generated realistic and diverse TOF MRA volumes of the Circle of Willis as indicated by visual analysis. The visual comparison of middle axial slices of the TOF MRA volumes and the 3D visualizations of the Circle of Willis anatomy between real and generated volumes is shown in Figure 4.

**Figure 4.**
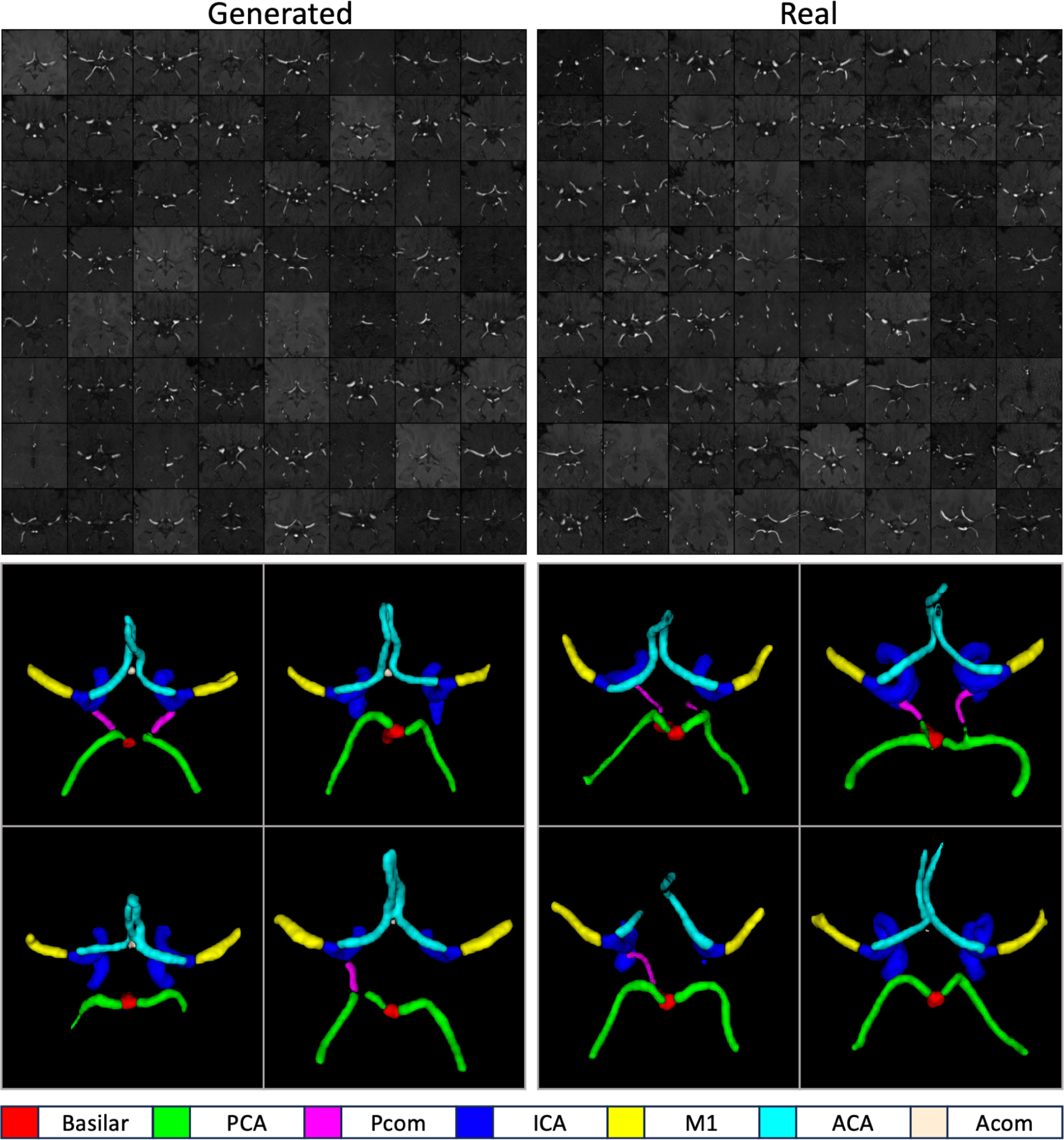
Overview of image quality and diversity of the generated images. Middle axial slices of random samples from the real and the generated datasets are shown (top section). Images are generated with the configuration Medium_v2 without the truncation trick. The segmentations used for 3D visualization of the Circle of Willis (bottom section) were generated using the teacher nnUnet model. Quantitative evaluation results: FID-axial 2D: 12.17, MedicalNet Distance-3D: 0.00078, AUC of the PRD: 0.9610.

Quantitative analysis was performed to select the data augmentation strategy and model complexity. The values of FID, MD and AUC-PRD were monitored during training (Figure 5). Best models from each run were selected by the lowest FID score among the three training steps with the lowest MD scores. The selected generator models were used to generate 1782 synthetic TOF MRA volumes matching the number of samples in the training set. The comparison of generated volumes against real volumes in the training set using FID, MD and AUC-PRD is shown in Table 3. The augmentation strategy was selected using the configuration *Medium_v1* as the baseline model. The model trained without any augmentations had the highest FID score, highest MD and lowest AUC-PRD and the training resulted in mode collapse as shown in Figure 5. All differentiable augmentations were combined with horizontal flip augmentations with a probability of 0.5. The combination of translation and cutout as differentiable augmentations had the highest AUC-PRD, outperformed the combination of translation, cutout and contrast with respect to MD and outperformed only translation with respect to FID (Table 3, Figure 5). Therefore, the translation and cutout augmentation was selected as the default differentiable augmentation strategy for model complexity analysis. The results of the model complexity analysis are summarized in Table 3. The configuration *Medium_v2* achieved the best performance (FID:12.17, MD:0.00078 and AUC-PRD: 0.9610). The configuration *Small* with the lowest capacity had inferior performance compared to more complex models in all metrics. Increasing the model capacity as in configuration *Large* did not lead to better performance.

**Figure 5.**
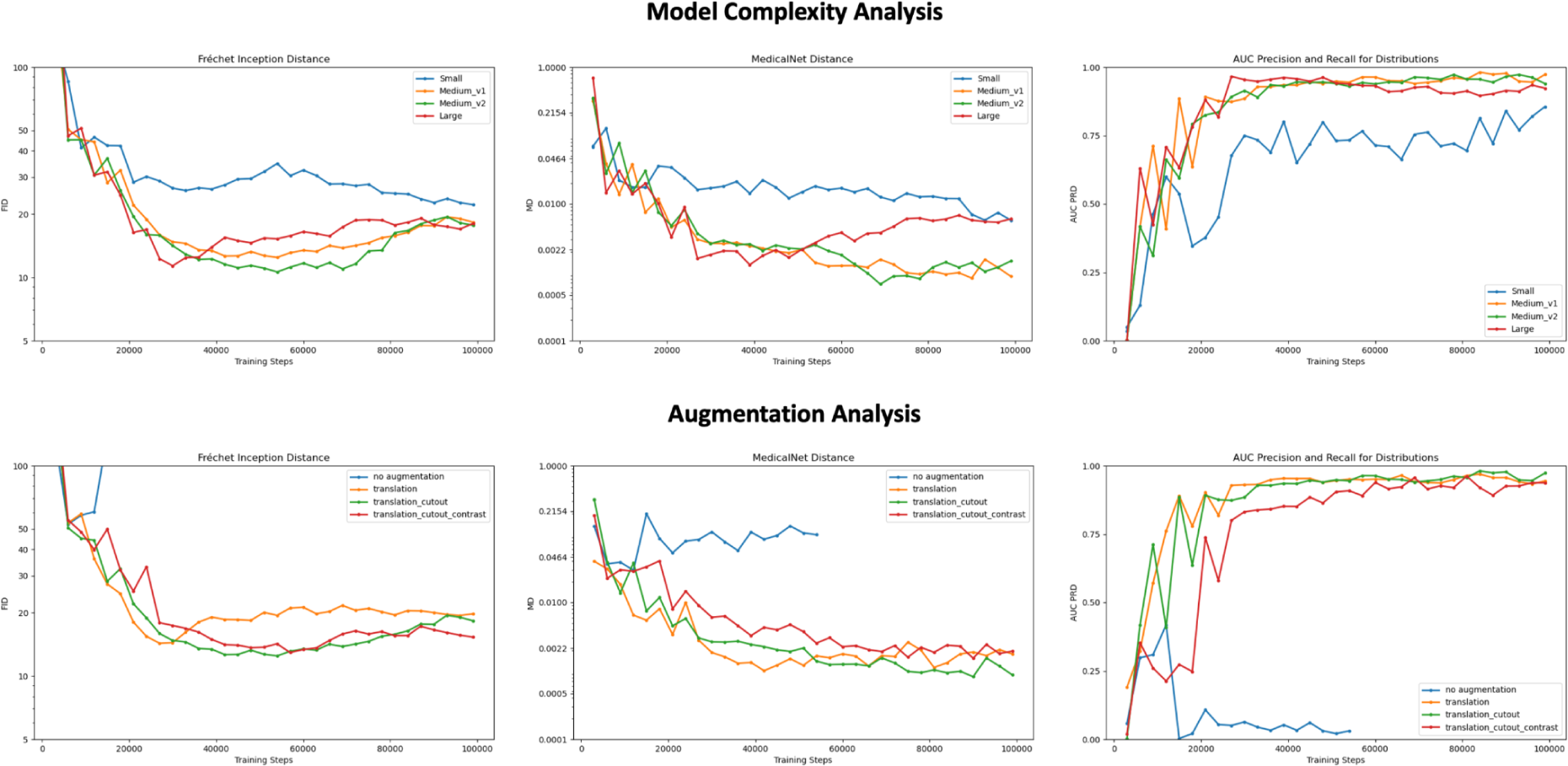
Model complexity and augmentation analysis. The use of differential augmentations improves training stability. The training for configuration “no augmentation” was stopped early due to the observed mode collapse. For the augmentation analysis the model configuration Medium_v1 was used. For the model complexity analysis, the translation_cutout strategy (green curve in the 2 row) was selected as the best augmentation strategy and used for all models. The configuration Medium_v2 was the best performing model of the model complexity analysis.

**Table 3.** Quantitative analysis of model complexity and augmentation strategy.

| Augmentation Analysis |  |  |  |
| --- | --- | --- | --- |
| Augmentations | FID | MD | AUC-PRD |
| No Augmentation | 54.11 | 0.04003 | 0.2891 |
| Translation + horizontal flip | 20.50 | **0.00086** | 0.9574 |
| Translation + cutout + horizontal flip | 17.14 | 0.00106 | **0.9687** |
| Translation + cutout + contrast + horizontal flip | **16.82** | 0.00233 | 0.9270 |
| Model Complexity Analysis |  |  |  |
| Model Complexity | FID | MD | AUC-PRD |
| Small | 23.05 | 0.00548 | 0.8263 |
| Medium_v1 | 17.14 | 0.00106 | **0.9687** |
| Medium_v2 | **12.17** | **0.00078** | 0.9610 |
| Large | 13.57 | 0.00209 | 0.9599 |

Quantitative evaluation on two random non-overlapping subsets of real TOF MRA volumes (n=891) resulted in FID of 1.54 MD of 0.00018 and AUC PRD of 0.9890. The comparison performed on two subsets (n=870) divided by datasets (Subset 1: OASIS, NITRC, CASILab vs Subset 2: Lausanne, IXI, ICBM) resulted in FID of 18.48 MD: 0.01334 and AUC PRD of 0.7926. Randomly selected patients from the OASIS dataset were excluded to match the number of cases in the two subsets in the second analysis.

In the visual turing test, the human rater reported to have considered the anatomy of CoW arteries, ophthalmic and anterior choroidal arteries, optic nerve, hypothalamus, pituitary gland, inferior horn of lateral ventricles, the Sylvian fissure as well as brain tissue details to distinguish between the images. The rater had an accuracy of 44 % in classifying the volumes as real or generated. A total of 100 volumes were evaluated (50 real, 50 generated). Out of 50 real volumes, 31 were identified correctly as real and 19 were falsely identified as generated. Out of 50 generated volumes, 13 were correctly identified as generated and 37 were falsely identified as real. The confusion matrix of the visual rating and respective example axial slices of TOF MRA volumes are shown in Figure 6.

**Figure 6.**
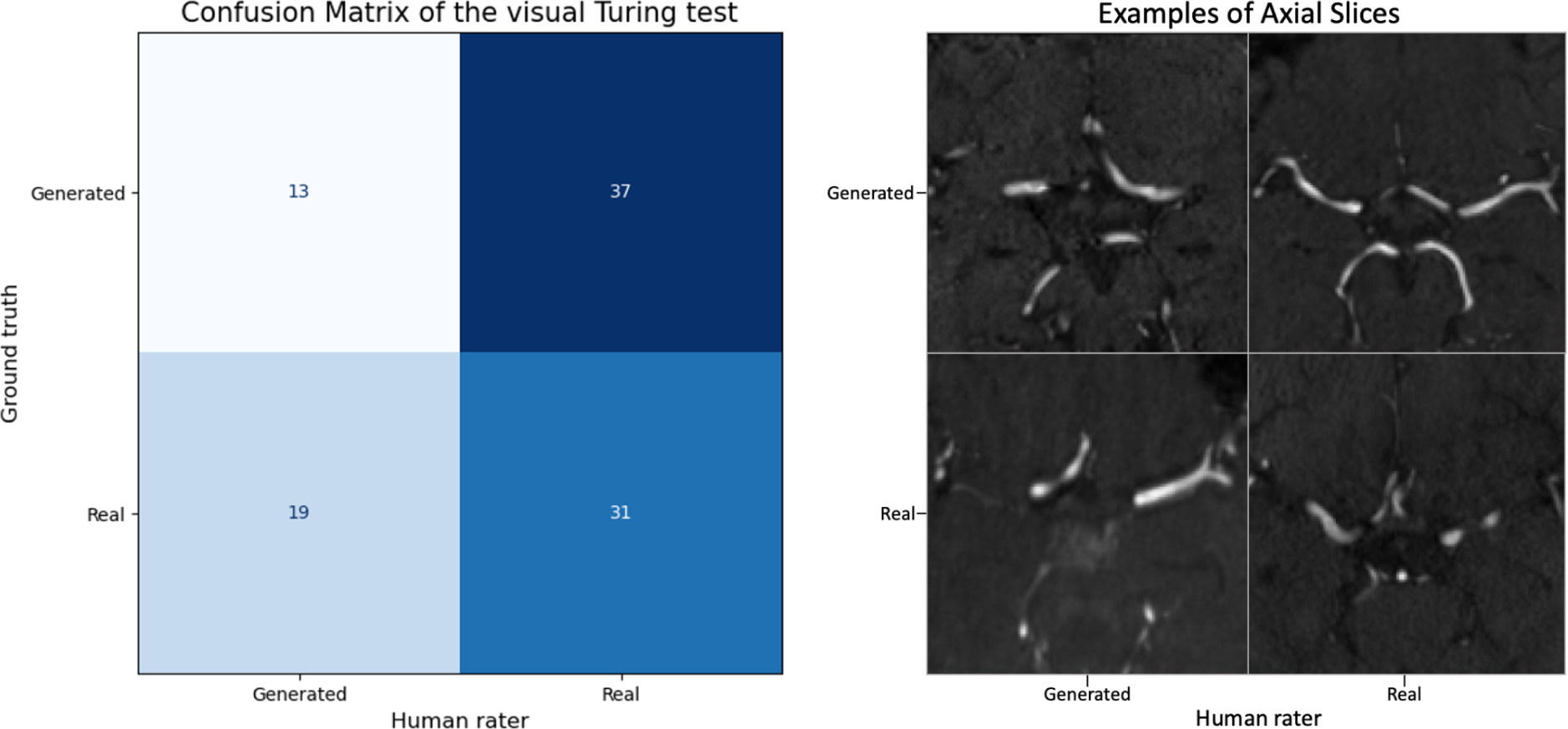
Visual Turing test results. The confusion matrix of the visual Turingon is shown (left). One example axial slice from each category (false positive, true positive, true negative, false negative) is shown (right).

Replica detection methodology based on the L2 distance could not identify any replicas in a random sample of 500 generated images. The visual analysis of a subset of 50 closest real images measured by the L2 distance was performed by the human rater and revealed no identical images.

In the following, the truncation trick, interpolation of style vectors and modeling of stochastic variations in images are demonstrated. First, the influence of the style scale parameter ψ on the diversity of generated images is shown in Figure 7. Decreasing the value of the style scale parameter images led to lower diversity and more homogenous contrast across the TOF MRA volumes. Second, the 3D StyleGAN model could generate a smooth transition of intermediate TOF MRA volumes when spherical interpolation between latent vectors was performed (Figure 8). Third, the stochastic variations in the images were modeled by changing the noise input of the model while keeping the style vector constant (Figure 9).

**Figure 7.**
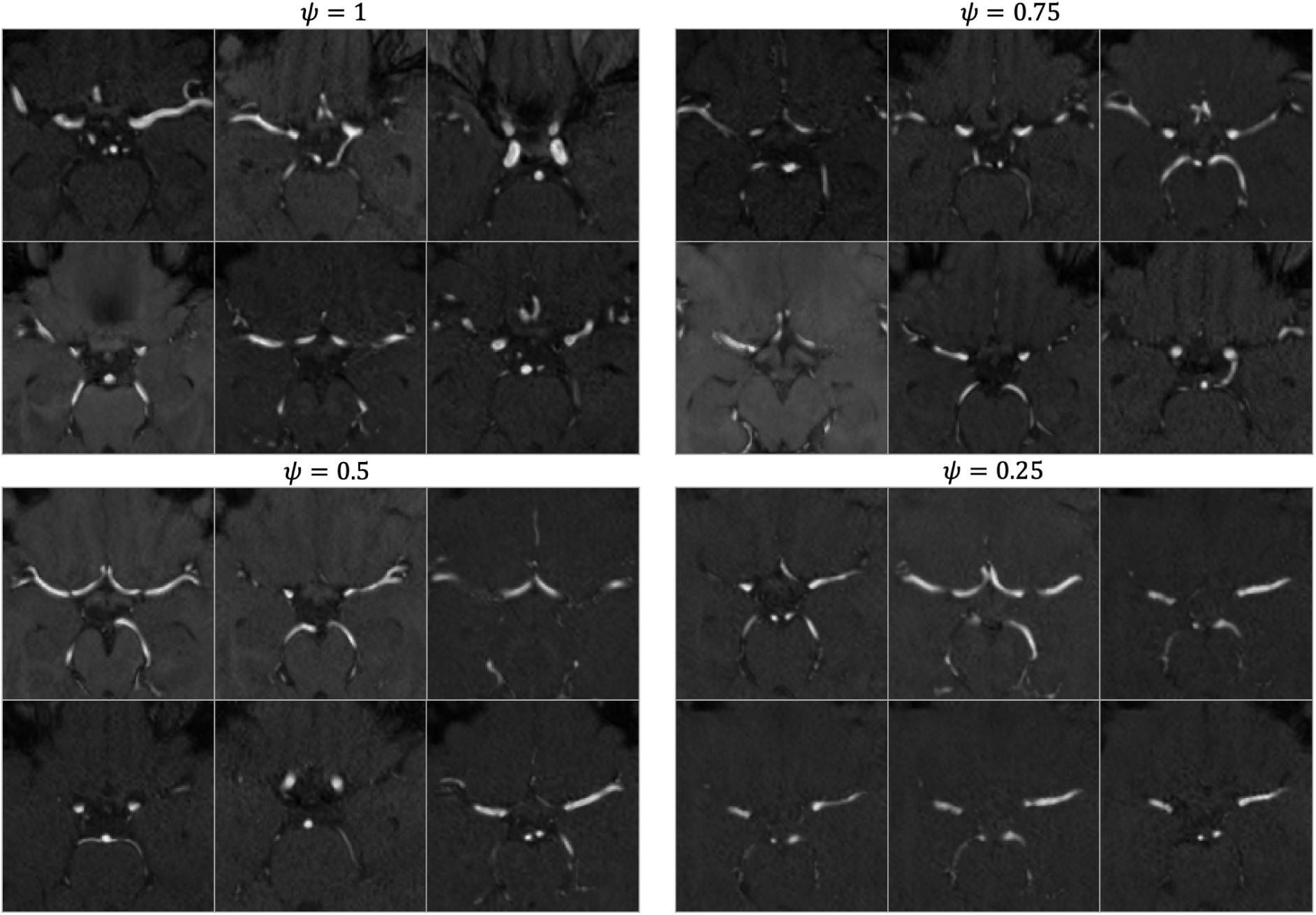
The influence of the style scale parameter ψ on the diversity of generated images. The truncation of the style vectors using lower values of ψ decreases the diversity of generated images. With lower ψ values, background details diminish, and the contrast across the images becomes more uniform, resulting in a set of images that appear more alike in anatomical features and overall visual impression (bottom left ψ=0.25).

**Figure 8.**
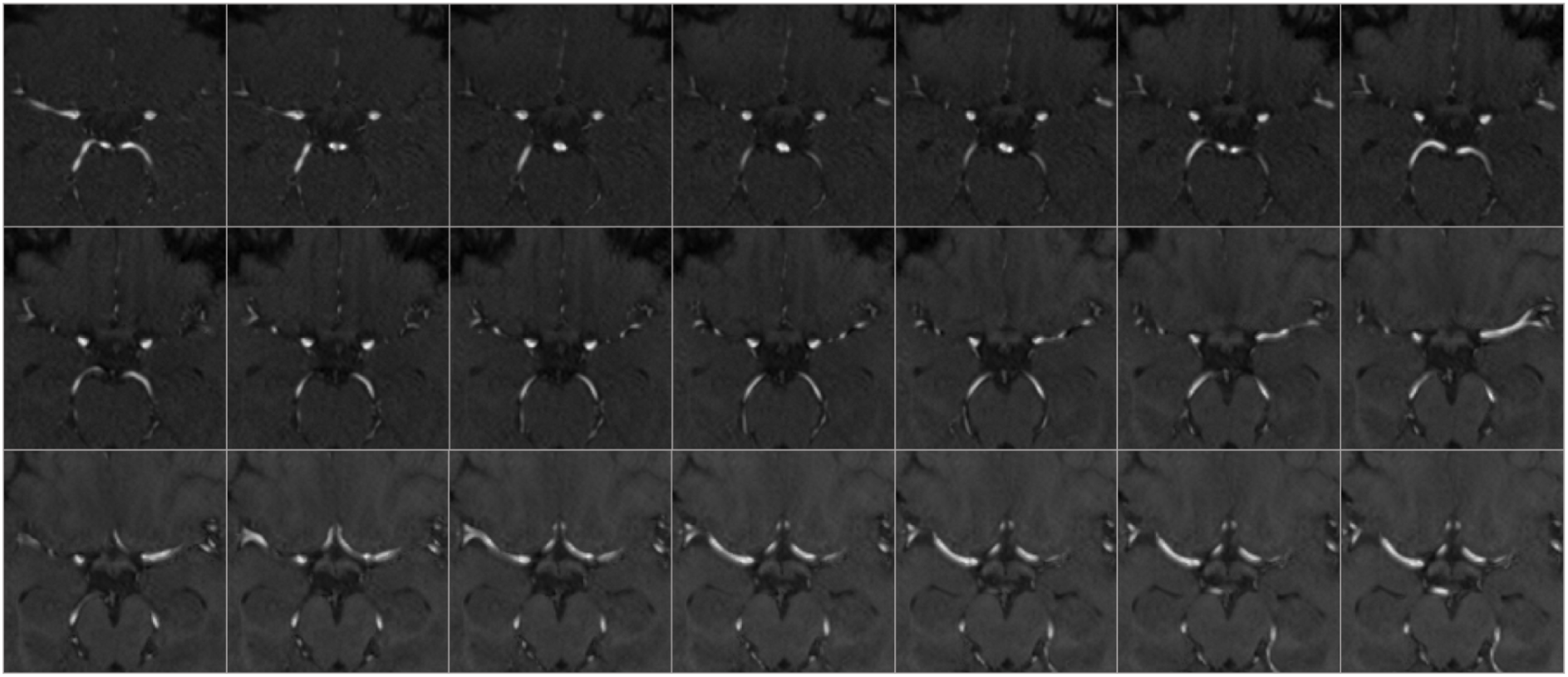
Image interpolation. From the image in the top left to the image in the bottom right a TOF MRA volume was generated for 20 intermediate steps using spherical interpolation between latent vectors. Smooth interpolation between images indicates that the model can generalize and shows limited overfitting. Truncation ψ was set to 0.75.

**Figure 9.**
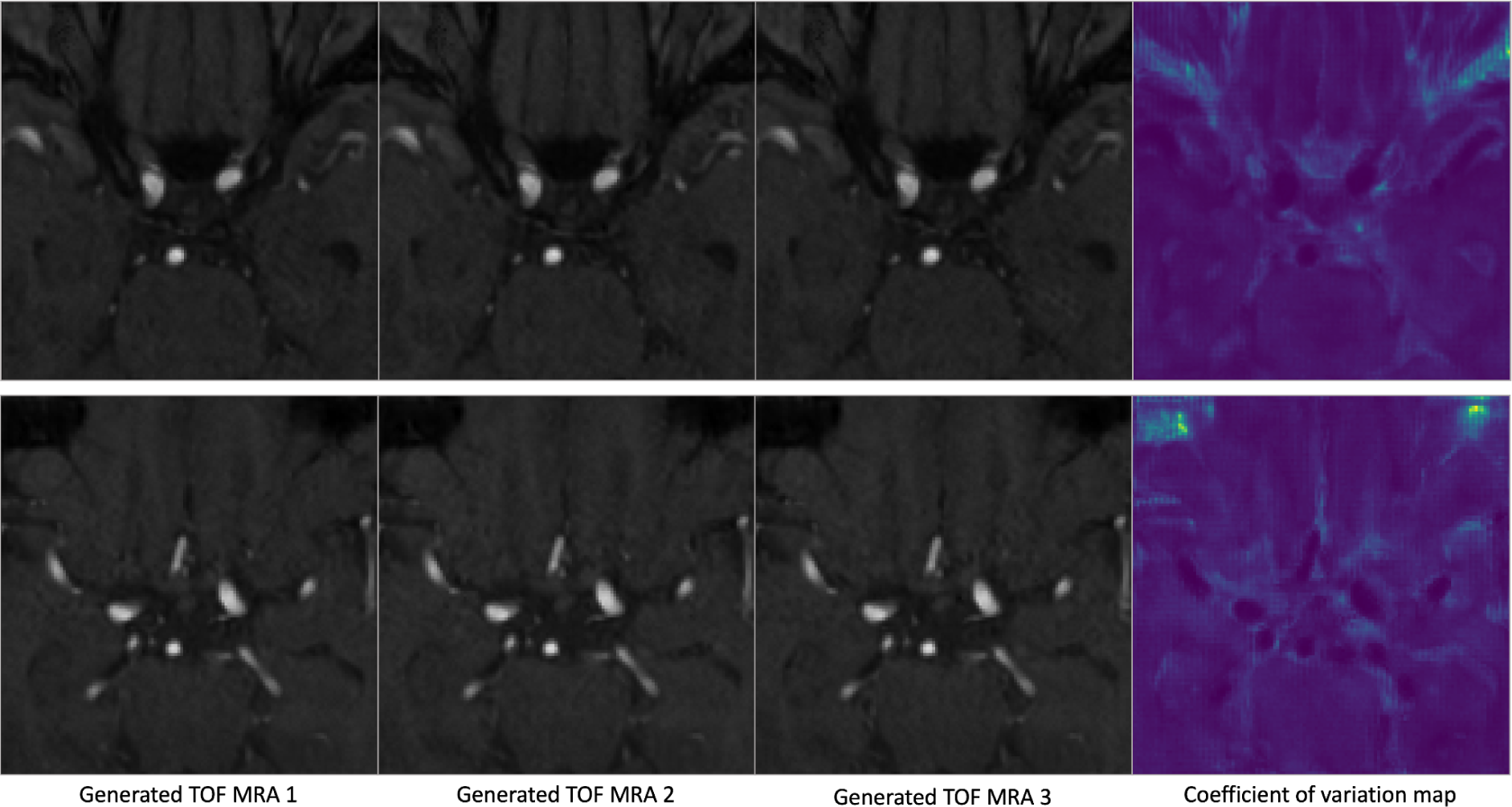
Generated images with stochastic variation. The noise input is varied while keeping the style vector the same across different TOF MRA volumes. The coefficient of variation map (right) shows the per voxel coefficient of variation over 100 generated sample volumes and highlights the areas that change with the noise input. While background details are controlled by the noise input, the vessel anatomy is controlled by the style vectors and remains unchanged.

For the downstream multiclass CoW artery segmentation, four student models trained on different training dataset compositions (50 real, 50 generated, 500 generated, 50 real + 50 generated) were evaluated on the test set of 60 patients from the TopCoW challenge. The quantitative results using the Dice coefficient and the Hausdorff Distance are summarized in Table 4. Increasing the number of generated volumes from 50 to 500 led to higher downstream vessel segmentation performance. Using 50 generated volumes in addition to 50 real patients as data augmentation in the training set did not lead to superior performance compared to the baseline teacher model trained on 50 real images. The downstream vessel segmentation performance in 3 example patients from the test set is visualized in Figure 10.

**Figure 10.**
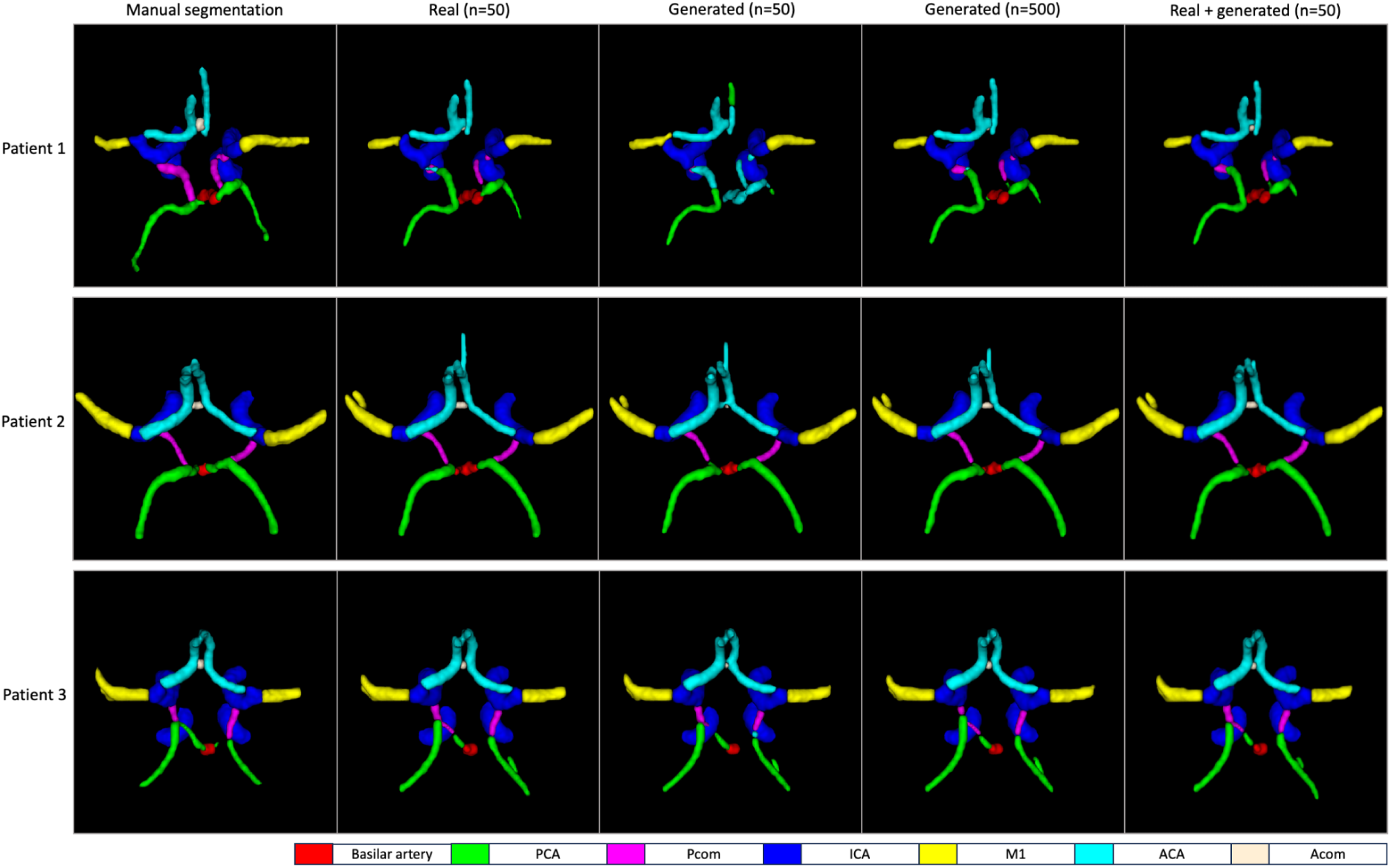
Visualization of downstream vessel segmentation performance using generated data. The student model trained on 50 generated volumes shows inferior performance on an unseen test patient compared to other models and misclassifies voxels of PCA as ACA and vice versa (Patient 1, third column). Notably Patient 1 has a more complex Circle of Willis variation compared to Patient 2. In Patient 2 and 3, all models demonstrate comparable performance, with the greatest performance variation being in the communication arteries Pcom and Acom. This observation is in alignment with the quantitative results from Table 4.

**Table 4.** Quantitative segmentation results. Mean values for the Dice coefficient and Hausdorff distance on the unseen test set are shown. The best mean value for each artery segment is highlighted in bold.

| Composition of Training set | ICA | M1 | ACA | Basilar | PCA | Acom | Pcom |
| --- | --- | --- | --- | --- | --- | --- | --- |
| Dice Coefficient (higher is better) |  |  |  |  |  |  |  |
| 50 Real | **0.909** | **0.789** | **0.834** | 0.871 | **0.798** | **0.498** | **0.589** |
| 50 Generated | 0.901 | 0.777 | 0.809 | 0.855 | 0.785 | 0.321 | 0.464 |
| 500 Generated | 0.907 | 0.785 | 0.829 | 0.870 | 0.795 | 0.419 | 0.554 |
| 50 Real + 50 Generated | 0.909 | 0.788 | 0.832 | **0.872** | 0.797 | 0.470 | 0.575 |
| Hausdorff Distance (lower is better) |  |  |  |  |  |  |  |
| 50 Real | 4.64 | 9.17 | **7.81** | 3.04 | 9.68 | 3.04 | **10.67** |
| 50 Generated | 6.42 | 11.15 | 11.03 | 3.05 | 10.09 | 3.46 | 14.00 |
| 500 Generated | 4.42 | **9.14** | 8.68 | **2.49** | **9.36** | 3.16 | 13.20 |
| 50 Real + 50 Generated | **4.33** | 10.23 | 7.86 | 2.59 | 10.00 | **3.00** | 11.94 |

## Discussion

We present an adaptation of the StyleGANv2 architecture tailored for the generative modeling of Circle of Willis anatomical variations. Generation of diverse TOF MRA volumes with high resolution and anatomical fidelity was possible with the proposed approach. Differentiable augmentations were of vital importance for 3D modeling with limited data. For downstream validation, we demonstrated the feasibility of training multiclass CoW artery segmentation models only on synthetic data and achieved comparable performance to a teacher model trained on real patient data. Synthetic TOF MRA data of the Circle of Willis can be utilized in a wide variety of clinical and medical imaging applications and can facilitate sharing of private medical patient data.

Generative modeling of 3D medical data poses unique challenges that need to be addressed in model development. Compared to 2D medical datasets, 3D datasets are more limited in size, data sharing can be more problematic and labeling in 3D is more labor intensive and costly. To tackle limited data and computational constraints in generative AI applications, previous works utilized modalities that are inherently 2D such as chest X-ray scans (Chambon et al., 2022) or histological images (Karras et al., 2020a) or extracted 2D slices/patches from 3D modalities such as MRI or CT (Kossen et al., 2021). Whereas hundreds of 2D slices or patches can be extracted from a 3D medical image, modeling in 3D significantly limits the number of training samples since each scan of a patient already includes multiple slices. 2D generative models are useful and practical in some downstream applications but lack the often necessary spatial consistency and anatomical context in many other medical applications. Applications such as hemodynamic modelling in cerebral arteries, large vessel occlusion detection algorithms, and aneurysm segmentation models require 3D medical data for model development. To address limited data scenarios in generative modeling, differentiable augmentations for the discriminator have been proposed by multiple research groups concurrently (Karras et al., 2020a; Tran et al., 2021; Zhao et al., 2020). We used the proposed differentiable augmentation strategy by Zhao et al. including translation and cutout together with horizontal flipping to increase generation performance in our limited medical data use-case. Our results suggest that without differentiable augmentations the generation performance of high quality and diverse data in the target resolution of 32×128×128 would be highly limited (Table 3). The model trained without any differential augmentations in Figure 5 resulted in mode collapse in the beginning of the training. Furthermore, to tackle computational limitations we trained the adapted StyleGAN on a cropped RO focusing only on the Circle of Willis and used mixed precision training. With this approach clinically common and acceptable image resolutions could be synthesized without sacrificing performance on our downstream task of multiclass CoW artery segmentation.

Generative models for medical imaging can be evaluated from various perspectives including fidelity (quality, resolution and anatomical realism), diversity (representation of variability in the real data distribution), utility (validation using a downstream clinical task) and privacy (data leakage) (Chen et al., 2021). For each generative modeling approach, the intended use case significantly defines model selection with respect to fidelity-diversity and privacy-utility trade-offs. In our study, fidelity and diversity of the generated images were equally important while we prioritized utility over privacy since our models are trained on already publicly available data.

Quantitative evaluation of image fidelity and diversity is rather challenging and controversial in medical imaging because most available metrics, such as FID, rely on feature extraction models pretrained on non-medical datasets containing natural images. The FID has been reported to be over-reliant on texture and is argued not to be directly transferable to medical images (Hong et al., 2021). Other works argue that FID aligns well with visual quality analyses (Woodland et al., 2022). In addition to FID, we therefore use the MedicalNet (Chen et al., 2019) for feature extraction before calculating the Fréchet Distance. Using the extracted feature embeddings by the MedicalNet we compute the area under the curve of the precision and recall for distributions curve similar to prior works (Subramaniam et al., 2022). In our use case, the quantitative results correlated well with visual impressions; the FID was used to assess the texture in axial slices, MD for the 3D consistency and overall quality and AUC-PRD for the diversity of the generated TOF MRA volumes.

Generative AI models are proposed to enable data sharing and aim to guarantee privacy by avoiding sharing real patient data. However one aspect that is neglected in some medical applications of generative AI is the assessment of replicas i.e. the memorization of real data from the training set (Pinaya et al., 2022). This phenomenon of generating replicas of real data is hypothesized to be a problem of overfitting due to limited data and can be more frequent in diffusion based architectures compared to generative adversarial networks (Akbar et al., 2023). Replica detection is especially important when no privacy preserving training methods such as differential privacy are used (Xie et al., 2018). Sharing of generator weights or generated data along with the publications might lead to restricted data being shared in an unnoticed way. Therefore, future works should implement a standardized replica detection method for generative AI applications in medical imaging.

To assess the utility of the generated data by the 3D StyleGAN model we assess the performance on the downstream task of multiclass CoW artery segmentation. The selection of this downstream task directly stems from the motivation of modeling the variations of the CoW and opens a path for a direct use case in clinical settings and medical imaging research. In this downstream task, we use a novel validation cycle based on the “teacher-student” model concept. In the initial step of the validation cycle, a teacher segmentation model trained on real data is used to provide pseudo-labels for the generated data. The ability of the teacher model to segment CoW arteries on generated TOF MRA data is a crucial finding, since it showcases that the quality and anatomical fidelity of generated samples are high enough to enable a satisfactory vessel segmentation result. In the second step, a student segmentation model trained on the pairs of generated data and pseudo-labels was able to segment unseen real TOF MRA volumes from the test set. The performance of the student model on real test data shows proper generalization and suggests that generated data retained the predictive properties of TOF MRA volumes for the CoW artery segmentation. Increasing the number of generated data for training of the vessel segmentation model improved segmentation performance, highlighting that increased dataset diversity and size is beneficial for CoW artery segmentation. Furthermore, the student model trained on 500 generated volumes produces high quality segmentations comparable to the model trained on 50 real volumes. These results are highly promising for future vessel segmentation approaches, since generated TOF MRA data and labels provided by either human experts or other models can be used to pretrain models or to perform on-the-fly data augmentation.

Generative models have important features that can be exploited for certain clinical and medical imaging applications. First, we explored image interpolation to enable a continuous transition between two generated images. Image interpolation has the potential to be used in different medical applications, for instance for disease progression modeling. Future works could train conditional generative models on large datasets including a specific pathology together with healthy controls. Interpolations between healthy images and late pathological images can provide important interim snapshots of a disease that may not be routinely captured due to lack of symptoms at an early asymptomatic stage of the disease. With this methodology, the growth of an aneurysm, the expansion of an intracranial hematoma, invasiveness and growth of a tumor or aging of an organ such as the brain can potentially be modeled, and respective datasets can be generated for specific downstream clinical or research applications. Second, the truncation trick allows control over the diversity and quality of the generated images. Lower truncation values scale the style vectors towards the mean style vector and potentially improve image quality while sacrificing the diversity of the generated samples. Thus, the truncation parameter can be tuned to fit the demands of a downstream use case to favor diversity or image quality. Rare variations of the CoW can be synthesized using higher values for the truncation parameter to create tailored datasets for rare variations of certain artery segments, such as the posterior communicating artery or the branching patterns of the middle cerebral artery. Third, the additional noise inputs in each layer in the StyleGAN architecture allow the modeling of stochastic elements in the generated medical images. Such stochastic variations can include background noise, different scanner qualities, or variations due to patient movement. Images with stochastic variations can be used as a form of data augmentation technique to improve robustness of downstream models towards adversarial attacks.

Our work has several limitations and implications for future works. First, although we used multiple open-source datasets we have utilized predominantly healthy patient data from a single imaging modality. We hypothesize that, given enough data, the results presented in this study can be translated to datasets containing pathologies such as aneurysms, strokes, or vascular malformations. Second, the 3D generative modeling is limited by computational resources with training times exceeding several days for training at clinically relevant resolutions and input sizes. To overcome this limitation, we trained the adapted StyleGAN by constraining the region of interest to only focus on the Circle of Willis and used mixed precision training. Due to computational limitations we did not perform exhaustive hyperparameter searches including the augmentation probability for the differentiable augmentation. Adaptive discriminator augmentations (Karras et al., 2020a) could be tested to improve performance in future works. Third, we have tested only the StyleGAN architecture for the modeling. More recent or efficient architectures such as Lightweight GAN (Liu et al., 2021) or diffusion models can potentially produce images with higher quality and diversity. Fourth, the dataset size of 1782 3D patient volumes is limited compared to non-medical datasets such as FFHQ (Karras et al., 2019) or medical datasets using 2D slices. We have limited our analysis to open-source data to allow for reproducibility and used techniques such as differentiable augmentations to alleviate this limitation. Fifth, in the downstream vessel segmentation task, we used pseudo-labels provided by the real model, effectively capping the performance of the student models to that of the real model, even when the generated data was used for data augmentation. Further studies could manually refine pseudo-labels provided by the teacher model to generate higher quality labels to potentially surpass the performance of models trained on real data. Finally, despite the replica detection methodology we employed, we cannot guarantee that our models are privacy preserving, i.e. data leakage in the sense of membership inference attacks cannot be ruled out where individual training data samples are recovered from our trained model. Therefore, we make only the code publicly available whereas our model weights will be made available upon reasonable request with assurance that the applying researcher has gained access to all the open access training datasets used in this study. This should let researchers reproduce our results and generate very similar synthetic datasets.

## Conclusion

In this study, we adapted the StyleGANv2 architecture to generate high-quality, high-resolution 3D CoW vessel data representative of inter-individual anatomical variations. Our approach demonstrates the feasibility of utilizing synthetic data for training multiclass CoW artery segmentation models, achieving comparable performance to models trained on real patient data. The utilization of differentiable augmentations, a cropped region-of-interest around the CoW, and mixed precision training helped to overcome the significant challenges of data scarcity and computational demands. Thus, our approach of generative modeling of 3D TOF MRA data paves the way for generalizable deep learning applications in cerebrovascular disease. In the future, the extension of the provided methodology to other medical imaging problems or modalities with inclusion of pathological datasets has the potential to advance the development of more robust models for clinical applications.

## Data Availability

open source TOF MRA repositories, please see Table 1 for data sources.

## Acknowledgements

Computation has been performed on the HPC for Research cluster of the Berlin Institute of Health.

The MR brain images from healthy volunteers used in this paper were collected and made available by the CASILab at The University of North Carolina at Chapel Hill and were distributed by the MIDAS Data Server at Kitware, Inc.

## Abbreviations

AUC-PRD: Area Under the Curve of the Precision and Recall Curve for Distributions
ACA: anterior cerebral artery
Acom: Anterior communicating artery
AMP: automated mixed precision
BA: Basilar artery
CoW: circle of Willis
FID: Fréchet Inception Distance
GANs: generative adversarial networks
ICA: Internal carotid artery
LVO: large vessel occlusion
MD: MedicalNet distance
M1: first segment of the middle cerebral artery
Pcom: Posterior communicating artery
P1: first segment of the posterior cerebral artery
ROI: Region of interest
ReLU: rectified linear unit
TTUR: Two Time-scale Update Rule
TOF MRA: time of flight magnetic resonance angiography

